# Meso-scale modeling of COVID-19 spatio-temporal outbreak dynamics in Germany

**DOI:** 10.1101/2020.06.10.20126771

**Authors:** A. Kergaßner, C. Burkhardt, D. Lippold, S. Nistler, M. Kergaßner, P. Steinmann, D. Budday, S. Budday

**Affiliations:** Department of Mechanical Engineering, Friedrich-Alexander-University Erlangen-Nürnberg, 91058 Erlangen, Germany; Department of Computer Science, Friedrich-Alexander-University Erlangen-Nürnberg, 91058 Erlangen, Germany

## Abstract

The COVID-19 pandemic has kept the world in suspense for the past months. In most federal countries such as Germany, locally varying conditions demand for state- or county-level decisions. However, this requires a deep understanding of the meso-scale outbreak dynamics between micro-scale agent models and macro-scale global models. Here, we introduce a reparameterized SIQRD network model that accounts for local political decisions to predict the spatio-temporal evolution of the pandemic in Germany at county and city resolution. Our optimized model reproduces state-wise cumulative infections and deaths as reported by the Robert-Koch Institute, and predicts development for individual counties at convincing accuracy. We demonstrate the dominating effect of local infection seeds, and identify effective measures to attenuate the rapid spread. Our model has great potential to support decision makers on a state and community politics level to individually strategize their best way forward.

## Introduction

Over the last months, we have observed the historic, global outbreak of the COrona VIrus Disease, COVID-19 (SARS-CoV-2). With the first official cases being reported in December 2019 in Wuhan, China [1], cases have since spread over the entire world, culminating in the world health organization (WHO) declaring it a global pandemic on March 11, 2020 [2].

Since then, each individual country had to find their own way to get the rapid spreading under control, to ‘flatten the curve’, and to avoid a breakdown of the healthcare system. Different strategies of shut down as well as travel and contact restrictions have been implemented in different countries and states, with more or less evidence for their success [3–5]. Now the great challenge is to slowly loosen restrictions but to avoid the feared second wave.

Germany has been given special attention during this pandemic. Firstly, its reported death counts were, especially initially, significantly lower than in neighboring countries such as Italy, Spain or France [6]. This gives rise to the question whether Germany could serve as an important example for successful strategies to mitigate the impact now and during future pandemics. Secondly, its federal structure has lead to different responses across its states to reduce human contact, to prevent further spreading, and now to slowly reopen the country and steadily go back to normal. And thirdly, the Robert-Koch Institute (RKI) provides locally highly resolved data on current cases that enables us to fit and test distributive models [7].

Besides the medicinal effort to understand the disease, numerous mathematical studies have focused on modeling the outbreak dynamics of COVID-19, predict its future course and provide scientific reasoning for political decisions. Typically, those epidemiology models follow the basic idea of compartmentalizing the entire population into different subgroups and modeling their coupled evolution with a set of ordinary differential equations (ODEs). The most basic of such models is the SIR model, with groups of susceptible, infectious, and recovered or removed people, dating back to the 1920s [8]. Overall, the course of a COVID-19 infection within such compartment models is quite well established by now. A susceptible is first exposed to the virus to become infected, before becoming infectious himself after some latency period. From here on, the infection may take various courses [9], ranging from no or mild symptoms for arguably the largest group of patients, to strong symptoms and patients who require hospitalization or even intensive care, before they recover or die from the disease. Severity mostly seems to depend on existing pre-conditions and general health, but also other reasons that have not yet been fully understood [10, 11]. The well-known SIR model has been extensively analyzed [12] and extended to finer compartments (see [13] for an earlier overview) that mimic the described course. Examples include the SEIR model with an <u>e</u>xposed group, the SEIRD model to separate truly recovered and <u>d</u>ead, an S(E)IQR model [14–16] that puts known infections into a quarantined group that does not infect others, or the MSEIR model [12] to include children with <u>m</u>other immunity, thus covering non-constant population sizes. Overall, these models have been abundantly applied to locally analyze COVID-19 outbreak dynamics in various countries, largely focusing on China [4, 17], Italy [14] and the United States (US) [18].

However, models to predict the temporal *and* spatial spreading of the virus have so far been rather limited. While agent-based models [19] successfully cover the high resolution end at the level of individual people and their movement, especially the intermediate to high resolution on a state or county level is understudied territory – even though this is exactly where many of the political decisions are being made. A variant of the SIRS model has previously been coupled to a reaction-diffusion model [20] to mathematically study cholera dynamics with partial differential equations (PDE). Colizza et al. have focused on the importance of the air travel network as a basis for global diffusion at a pandemic outbreak [21]. Following this strategy, Ellen Kuhl’s group at Stanford have coupled an air travel network to the SEIR model to understand spatial spreading in China, the US [18] and across Europe [22]. However, with air travel being almost entirely cancelled, we are in need of short-to mid-range network models, such as [23]. While the global epidemic and mobility (GLEAM) model [24] includes air travel as the major source of wide-range disease spreading, it also models more localized commuting patterns that correspond well to traffic data in Germany, among other countries. The model has explained a great deal of COVID-19 spreading in mainland China [3].

As suggested by multiple previous studies [25, 26], mildly or asymptomatic carriers account for the major share of new infections, and the large number of hidden infections facilitated global spreading [27]. Thus, here we model the spatio-temporal outbreak dynamics of COVID-19 in Germany with an SIQRD model that specifically distinguishes between the hidden infectious I group and a Q group that holds people with known, quarantined infections that, consequently, do not infect others anymore. Since Germany was several weeks behind China and Italy with several global travel restrictions already being in place, we couple the SIQRD model to the GLEAM mobility network to model short-range and intra-country interactions essential to locally resolve the evolution of the COVID-19 pandemic.

## Results

### The basic SIQRD model

Fig. 1 demonstrates how the overall COVID-19 cases in Germany would have evolved at initially observed exponential growth without any political interventions – based on the optimized SIQRD model with the model structure and dynamics displayed in the schematic in Fig. 1 (parameters in suppl. table S1). The model estimates that Germany could have suffered nearly half a million dead. Note that the total number of currently infected corresponds to I+Q. Our model yields an initial reproduction number of *R*_0_ ≈4.61 in early March, consistent with previous studies [11, 28].

**Fig 1.**
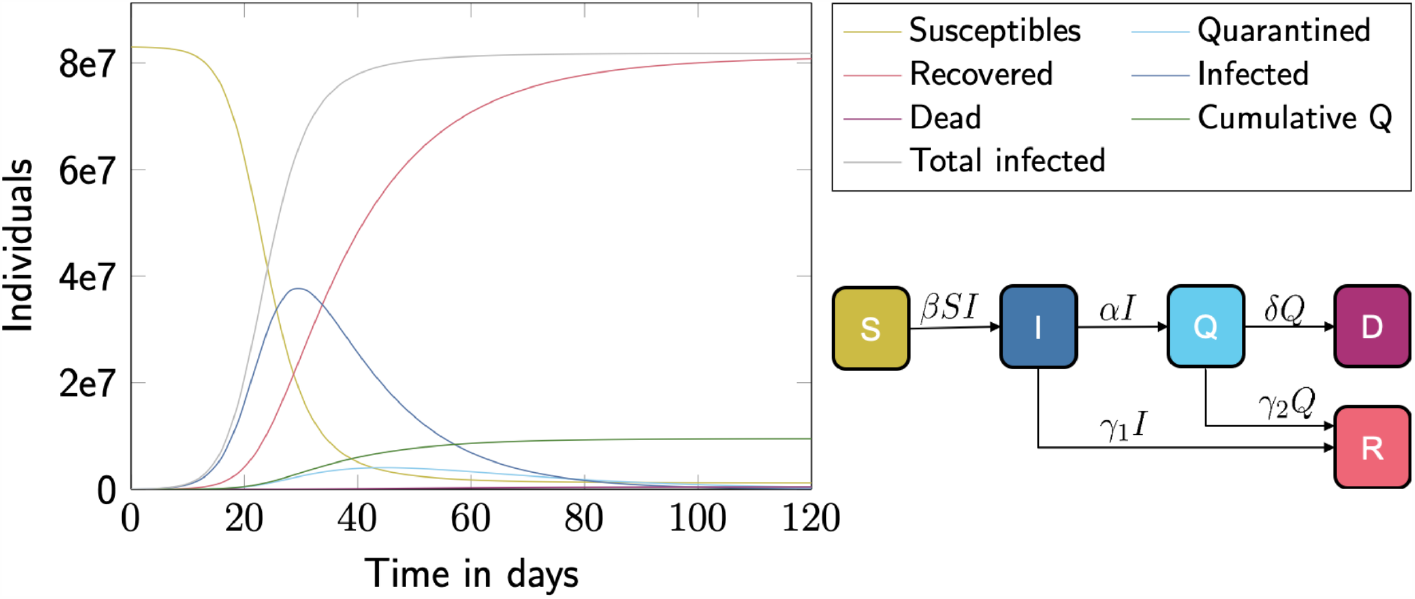
The basic SIQRD model. Unrestrained evolution of the pandemic in Germany based on the basic SIQRD model (schematic lower right) with parameters fit to the initial exponential growth (left).

As Germany claimed high testing rates from the beginning on, we refrain from including a time-dependent *α*, denoting the rate at which people transition from the infectious group I to the tested, confirmed, and quarantined group Q. Instead, for a more natural interpretation of the transition rates, we replace *α* and *δ* – the rate at which diseased individuals in Q die and enter the group D – by introducing the dark figure *ω* and the true mortality *µ*, also referred to as infection fatality rate (IFR), and obtain

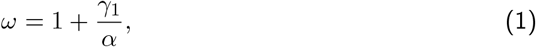

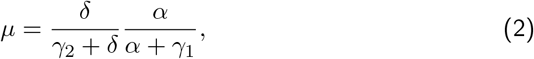

where *γ*_1_ and *γ*_2_ quantify the recovery rates from hidden (I) and quarantined (Q) infections, respectively. The reparameterization allows us to explicitly identify the role of the dark figure in COVID-19 spreading. The measurable ratio D / Q, usually referred to as case fatality rate (CFR), will eventually approach the (constant) product *ωµ*, demonstrating the inherent coupling of the two parameters. However, during the course of the pandemic, CFR is not constant due to the time delay between D and Q, a potentially varying dark figure *ω*, fluctuating testing capacities, and a disparate mortality across the age structure of infected people [29].

The pivotal group for the disease dynamics are the infectious people I, while infection numbers reported by institutions such as the RKI correspond to the daily or cumulative influx to Q. It is obvious from the model that reducing the number of infectious people in I is achievable by reducing the infection rate *β*, the size of S [4] and/or the size of I itself. The latter is, on the one hand, indicated by a faster transition rate *α* from I to Q, e.g., by increased testing: Once people have been positively tested, they will quarantine and likely not infect others any more. On the other hand, a faster transition rate *γ*_1_ will reduce people in I, e.g., by general protective and hygienic measures: If an individual is exposed to a lower viral load, symptoms and the general course of the disease may be milder [30].

### Accounting for state-wise political measures and testing

To capture the influence of political decisions and restrictions on the disease dynamics, we have included three reduction factors for the infection rate *β*_*red*,1−3_ representing the major restrictions of 1) cancelling large events, 2) school closings, and 3) contact restrictions. We model each of them as a single constant factor for all of Germany, but consider their federal-state-dependent starting date. Taken together, these political decisions, implemented at the right time, successfully slowed down the spread by reducing the number of daily new infections – the influx to Q – compared to the unrestrained evolution displayed in Fig. 1, and have thus prevented an overburdened health care system, as seen in Bergamo or New York City.

A much less tangible group than the detected infections in Q are the hidden infections in I. By assuming a Germany-wide identical mortality of *µ* = 0.006 [9], we obtain estimates on state-wise dark figures *ω*_*i*_ (Fig. 2a) by fitting to the individually reported death tolls, with *(ω*_*i*_) ≈ 8.64 (see Methods, suppl. table S1). Interestingly, we observe a fair, negative correlation (Pearson coefficient *r*_*P*_ = −0.62, *R*_2_ = 0.38 *p <* 0.0085) between state-wise *ω*_*i*_ and performed per-capita tests, which varied significantly from about 0.02% of the population in Saarland to about 1.44% in Berlin (test numbers from April 24, suppl. table S1 [31]).

**Fig 2.**
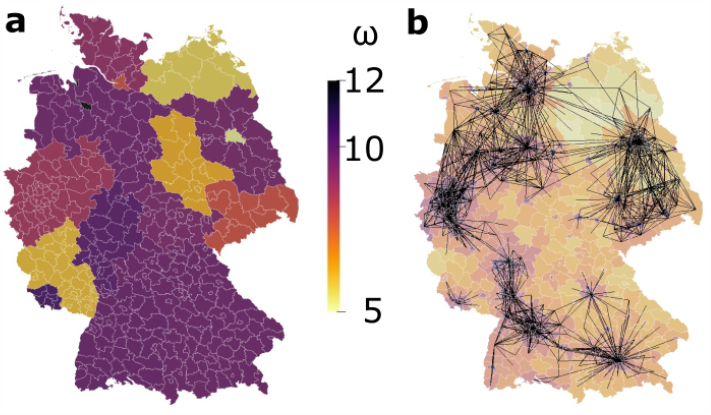
Federal-state-wise dark figure and cross-county interactions. (**a**) State-wise estimated dark figures following from a Germany-wide constant mortality. (**b**) Major connectors (lines) between counties of the network mobility model to predict cross-county infections in the spatial SIQRD model. County color varies from yellow to dark purple with population density.

### The spatially resolved SIQRD model

To obtain spatially resolved outbreak predictions, we combined the refined SIQRD model with an adapted short- and mid-range mobility network from GLEAM [32]. The latter has been shown to well capture commuting patterns in Germany, as described in [24]. It accounts for the interaction between different cities and counties throughout Germany, as illustrated in Fig. 2b. To adequately represent cross-county infections considering severely altered mobility patterns in times of the pandemic, we introduce state-wise cross-county multipliers *β*_cc,*i*_, *i* = 1, …, 16 to modulate the exponential cross-county term.

With state-wise identified dark figures *ω*_*i*_ (Fig. 2a) and otherwise estimated parameters *µ* = 0.006, *γ*_1_ = 0.067, *γ*_2_ = 0.04 (see Methods), we obtain optimized state-wise infection rates *β*_*i*_ and cross-county weights *β*_cc,*i*_, as well as Germany-wide contact reduction factors *β*_red,1−3_. Importantly, our preliminary investigations had shown that it is not sufficient to provide a single *β* valid in entire Germany, even with state-wise dark figures *ω*_*i*_. It is, therefore, key to calibrate infection rates *β*_*i*_ differing between federal states. We further note that current data do not allow us to clearly distinguish three independent reduction factors. Especially, effective dates of *β*_red,2_ (school closings) and *β*_red,3_ (contact restrictions) are close by in time, leading to similarly good fits with various pairs of the two. To prevent over-fitting, we thus set *β*_red,2_ = 1 and use one combined reduction factor for school closings and contact restrictions for our spatial simulations instead, optimized to *β*_red,1_ = 0.378 and *β*_red,3_ = 0.189 (see Methods).

Another important component of spatially resolved predictions is the choice of appropriate initial conditions. Besides the mere size of *I*_0_, the initial infectious population, we also needed to specify its spatial distribution at the start date of the county-wise simulation, March 2. Due to several days delay between the outbreak in different federal states and, naturally, their different population size, we selected the RKI reported distribution on March 16 and scaled its magnitude down to obtain the overall number as determined on March 2, state-wise amounting to *(I*_0,*i*_) = 2679 (see Methods, suppl. table S1).

Fig. 3 demonstrates that the optimized spatially resolved SIQRD model with 401 network nodes representing each county of Germany well predicts the cumulative confirmed cases in each of its federal states from March 2 until April 25. For cumulative infection data reported by the RKI [7], we find astonishing agreement on the temporal evolution (all *R*_2_ *>* 0.96). Some states, most noticeably Bremen, show rather irregular numbers of daily new infections and slight deviations from the model, which we attribute to its low testing rate (second lowest in Germany), a corresponding high dark figure (Fig. 2a), and the overall very low case counts. The model slightly overestimates the number of deaths in the initial period but well matches the overall number, the only dead count included in the fitting procedure (Methods). This can be attributed to a strong age-dependence in mortality [10, 11] and the fact that the spread of COVID-19 in Germany started off not least due to – mainly younger – infected returnees from skiing vacations in Italy and Austria [29]. Later on, more and more elderly people caught the disease, which then lead to an overall stronger death toll around the early midst of April, including devastating death rates in several nursing homes.

**Fig 3.**
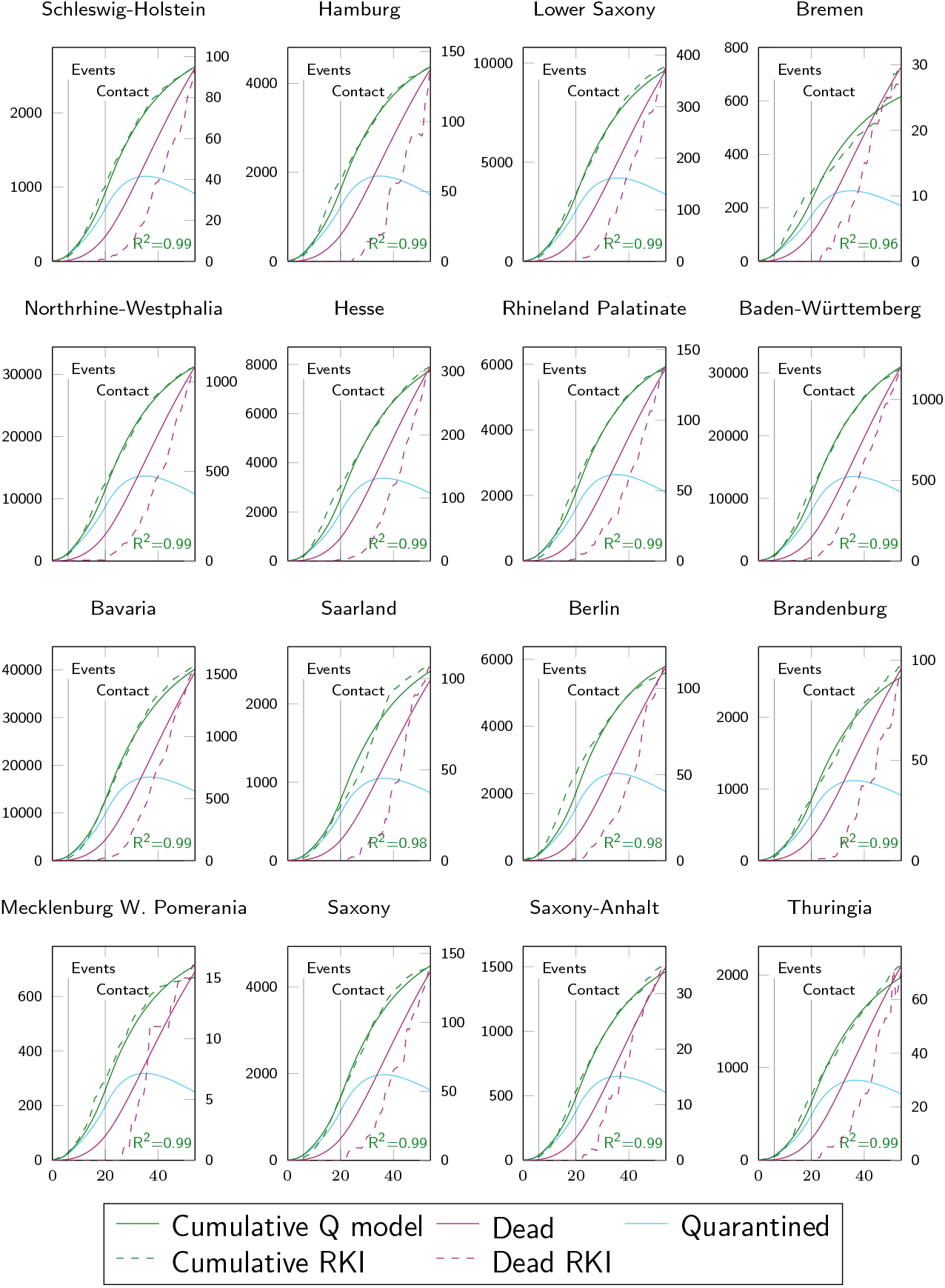
State-wise predictions of the spatially resolved SIQRD model. The evolution of cumulative quarantined infections Q (left axis, green; current Q in teal) and dead count on last day (right axis, purple) were state-wise fitted to RKI data (dashed) from March 2 to April 25 (*x* in days since March 2). Vertical lines denote changes in *β. R*^2^ indicates goodness of fit.

Fig. 4a and b display federal-state-wise *β*_*i*_ and *β*_cc,*i*_. On average, the intra-county infection rates *β*_*i*_ are about half as large as for the basic SIQRD models [14], leading us to believe that about half of infections occur through cross-county interactions. We observe an opposite trend between *β*_*i*_ and *β*_cc,*i*_, suggesting a trade-off between intra- and inter-state contact. Remarkably, northern (touristy) regions obtain higher inter-state contact rates, suggesting they observed relevant inflow from other states. Furthermore, highly populated states, most notably the city-states Berlin, Hamburg and Bremen (isolated black dots in Fig. 4b), but also Northrhine-Westphalia (NW), Baden-Württemberg (BW), and Bavaria (BY), seem slightly over-represented in the network and thus receive smaller *β*_cc,*i*_. This is to be expected, as densely populated areas typically observe more mobility and commuter traffic, which was more severely reduced when the pandemic hit.

**Fig 4.**
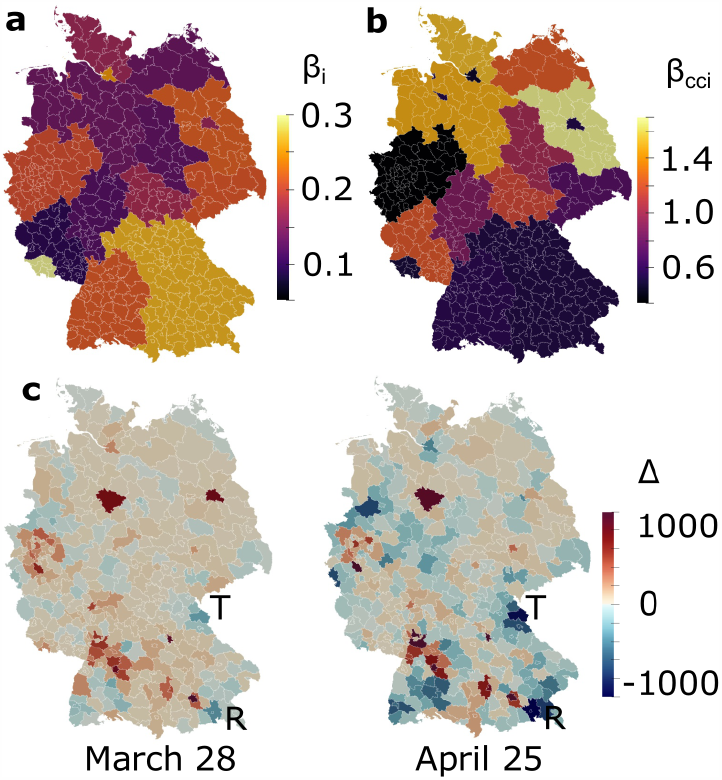
Optimized model parameters and validation. Illustration of state-wise optimized values for *β*_*i*_ (**a**) and *β*_cc,*i*_ (**b**) for *i* = 1, …, 16. (**c**) County-level evaluation of cumulative quarantined infections Q on two randomly selected dates, showing the difference of infections SIQRD −RKI, increasing from blue to red, with indicated counties Tirschenreuth (T) and Rosenheim (R). Color scale fixed to [-1000,1000] from [-305,3206] on March 28 and [-1197,4896] on April 25 for comparability and better contrast.

### County-wise predictions

We then analyzed how well the federal-state-wise fitted model represented the infections on a county level. Fig. 4c shows the difference of cumulative entries in Q between our model predictions and RKI reported numbers on two dates in April, without further county-wise fitting or optimization. On both dates, we find a strong, significant correlation of the spatial distribution (March 28: *R*_2_ = 0.84, *p <* 1*e*−16; April 25: *R*_2_ = 0.81, *p <* 1*e* −16), demonstrating an overall high level of accuracy of our meso-scale model. On the earlier date in March, larger differences are almost exclusive to Hannover and Berlin, two densely populated cities rather distant from major initial hubs. One month later, we observe slightly more deviations overall. Not surprisingly, they mostly occur in BY, BW and NW, the three most populous states with highest infection numbers overall, putting the differences into perspective.

Comparing per-capita cumulative infections on March 28 (April 25), we find that, despite a lowered correlation, 79% (57%) of counties differ less than 50 per 100000 inhabitants, which is the politically imposed threshold for daily new infections to re-implement restrictive measures in most German states. We also observe some rural areas that suffered from more infections than predicted by our model. The most prominent examples are the indicated counties Rosenheim (R) and Tirschenreuth (T). Per-capita infections in the hot-spot city Mitterteich in Tirschenreuth temporally surpassed the numbers in New York City [6] and one of the most stringent curfews was put in place to contain the virus spread [33].

Following this validation, we used our model to obtain a complete spatio-temporal timeline of the COVID-19 spreading. Fig. 5a and supplementary movie S1 show the predicted spatial distribution of all infections, i.e., combined entries of I *and* Q, evolving from early March until early June at the resolution of individual counties, assuming the contact reduction factors stay in place. The snapshots nicely capture the more severe course of the pandemic in the Southern and Western states, while Eastern Germany was less affected.

**Fig 5.**
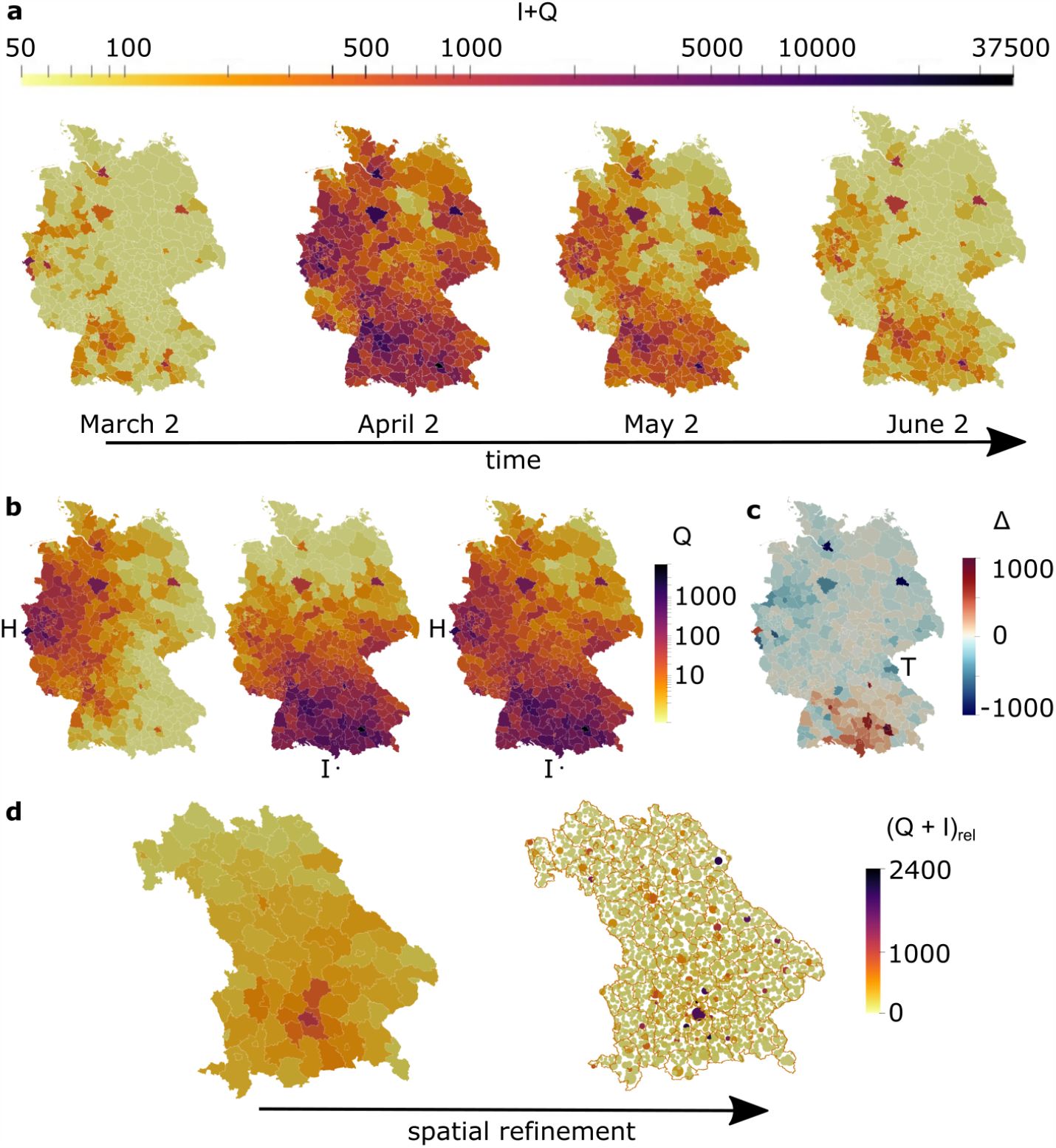
Model predictions. (**a**) Temporal snapshots of the epidemic spread across Germany, continuing with the identified reduced infection rates *β*_*i*_. (**b**) Scenario comparison on March 28 close to the epidemic peak, with seeds spreading from Heinsberg (H, left), Ischgl (I, center) and both cities (H+I, right). (**c**) Difference plot of cumulative infections Q between our simulation with seeds from Ischgl and Heinsberg (panel B, right) vs RKI data on March 28. Color scale cropped from [-2230,6550] to [-1000,1000] for better contrast. (**d**) Refined predictions on April 11 in Bavaria at county vs. city (circles) resolution. Scale shows cumulative infections per 100,000 inhabitants.

### The effect of seeds

The spreading of COVID-19 in Germany has allegedly evolved from two major hubs: (1) a carnival event in the county Heinsberg (H) in the Ruhr area in Western Germany, and (2) returnees from skiing holidays in Northern Italy and Austria, with a large share tracing back to Ischgl (I). With the aid of our spatially resolved model, we investigated how these sources may have affected the spreading throughout Germany.

Fig. 5b shows the distributions of confirmed, quarantined infections Q resulting from initial outbreaks in Heinsberg (left) and Ischgl (center) only, as well as the distribution resulting from the combination of both (right), demonstrating the balance of local and far-reached infections with our model. Red hot-spots appear in nearly all major urban areas across Germany, but rural spreading occurs much more in areas closer to the seeds. When evaluating differences of infections from their combined spreading to RKI data (Fig. 5c) near the German peak on March 28, we find that, despite increased differences very close to the Southern border, overall state-wise distribution in BY is near identical in quality (*R*_2_ = 0.89 in both cases). This demonstrates the dominating influence of returnees from skiing holidays in Italy and Austria represented by the Ischgl seed for the Germany-wide initiation of the spread. Tirschenreuth (T), however, seems to have suffered from their very own, less documented super-spreading event. On the other hand, the distribution in NW differs more to the data (*R*_2_ from 0.79 to 0.64). We observe several underrepresented counties, suggesting that cases from Heinsberg alone spread less, similar to the localized situation in Tirschenreuth. The overall similarity between Fig. 5c and Fig. 4c, as well as Fig. 5a and Fig. 5b does confirm, however, that the virus indeed spread from the Southern and Western states of Germany, with Ischgl and Heinsberg as two major representative seeds.

### Meso-to micro-scale

It is possible to further refine the resolution of the spatial SIQRD network model to account for individual cities. Fig. 5d compares the county-wise (left) with the even more refined city-wise model (right) in Bavaria on April 11. While the numbers of the whole county are controlled by larger cities, the more refined model also captures lower (per-capita) infections in small communities. The increased resolution may provide valuable information for local decision makers, especially in more rural areas where the epidemic course is not as much controlled by the closest major city.

## Discussion

We have shown that a spatially resolved SIQRD model can well explain and predict the temporal *and* spatial outbreak dynamics of COVID-19 in Germany. The reparameterized model specifically includes undetected, hidden infections as a separate compartment, revealing a direct coupling between mortality, testing efforts and the dark figure. Our systematic refinement from Germany-wide to spatially resolved county-level predictions has revealed that we require different values for dark figures *ω*_*i*_, infection rates *β*_*i*_ and cross-county weights *β*_cc,*i*_ in each federal state of Germany to accurately capture the spreading of COVID-19 from March until May 2020. At first, this is quite surprising considering that various other studies with single-node, country-wide models have predicted single infection rates *β* that are quite similar for different countries, such as Germany, France or Spain [11, 14]. However, this can be attributed to their low spatial resolution and high infection numbers, which average out any spatio-temporal fluctuations. Higher-resolution information as presented here thus comes at the cost of more complex model requirements.

Differences in *ω*_*i*_ can in part be attributed to variable testing activities. It is important to note, however, that the varying dark figure alone is not enough to account for the different outbreak dynamics in the federal states of Germany. Rather, there seems to be a non-negligible influence of habits and mentality that drive different infection rates *β*_*i*_, together with random factors and local super-spreader events such as the carnival celebrations in Heinsberg [30]. We observe an opposite trend between *β*_*i*_ and *β*_cc,*i*_ (Fig. 4a and b), suggesting that some states (mostly Northern and distant from initial seeds) received more infections from neighboring states, while states close to epidemic seeds suffered more from localized infections. Generally, the adapted mobility network tended to overestimate cross-county terms in densely populated areas, where the pandemic seemed to have a larger reduction effect on typically observed traffic patterns, manifested by smaller weights *β*_cc,*i*_.

It has become clear from our analysis that the data we currently have at our disposal makes it impossible to provide ‘true’ parameter sets that uniquely describe the evolution of the pandemic. However, despite the deduced interdependence of mortality *µ* and dark figure *ω*, the relationship to testing activities holds regardless, underlining the importance of broad, fast testing. In addition, increased (antibody) testing can help strengthen our confidence bounds on *ω* and *µ* in the future. Similar relationships exist for the politically induced reduction factors *β*_red_, where data only allowed us to distinguish two independently.

Our spatially resolved model can predict the temporal evolution of infections on a county level at convincing accuracy, and even extends to individual cities (Fig. 5d). It nicely captures the fact that the probability for new incoming infections and higher spreading is generally larger in densely populated urban environments. However, we have also seen a few rather rural counties with high infection numbers that were much less hit in our predictions, e.g., the county Tirschenreuth in Eastern Bavaria (Fig. 4c). We postulate that such locally over-proportionate case counts can be attributed to rather random super-spreading events, which may pop up anytime and can easily be included in our model, but are hard to predict in advance.

Exploiting our county-level resolution, we were able to infer the effect of infections stemming from selected seeds, such as two major hubs for Germany, Heinsberg and returning travellers from Ischgl in Austria. Our model demonstrates how the outbreak dynamics in Germany were initially driven by these two major seeds and spread from there throughout the rest of the country (Fig. 5b). Nevertheless, from our difference analysis we found that Heinsberg itself was far more contained than Ischgl (Fig. 5b).Taken together, these observations corroborate that refraining from traveling and large events are two key interventions that can effectively attenuate the spreading of infectious diseases such as COVID-19.

The presented model has certain limitations that we aim to address in the future. One drawback of all SIR-type modeling approaches is that they hardly account for the various courses of disease: in such rate-dependent models, some appear as infinitely long infectious. To prevent this issue from significantly affecting our optimized parameters, we only considered the latest dead count in our fitting procedure (see Methods). Still, we plan to adapt our model to integrate detailed information on specific courses of disease within a memory-based or delayed ODE, as introduced e.g. in [34].

Most noticeable deviations of our model predictions on a state level occurred in Bremen, a city-state with overall very low infection numbers. Despite a high dark figure and concomitant uncertainty, on the city level our quasi-continuum modeling approach and the underlying exponential growth seem to approach their validity limit, while stochastic effects start to become more important. Whereas SIR-type compartment models may capture the spread on a macro- and meso-scale level, at very low infection numbers or high spatial resolution, individual agent-based models [19, 35] are required to accurately predict the course of the epidemic. It is noteworthy, though, that current agent-based models may scale up to ≈ 50.000 agents, leaving quite a gap to meso-scale models like ours. We will investigate how coupling both types of methods in a multi-scale model can close this gap in the future. Similarly, explicitly integrating uncertainty via stochastic models [36] may help to further improve model predictions at high spatial resolution and low to medium infection numbers, potentially providing insights into optimal strategies for political action.

Overall, our refined predictions could provide a trustworthy rationale to elaborate community-wise reopening and closing strategies, safely plan the occupation and need of hospital beds, and inform on optimal distribution strategies of vaccination and/or antibody tests once available. The optimized model can be directly adopted to estimate the effects of loosened restrictions, potential new seeds, or other influencing factors on the resolution of individual cities. It can thus be a valuable tool to support (political) decision makers to appropriately react to future developments of the COVID-19 situation and expediently avoid a second wave.

## Methods

We model the spatio-temporal outbreak dynamics of COVID-19 in Germany with an SIQRD model, coupled to a network model that allows for spatially distributed cross-county infections. We start out with the description of our basic compartment model that mainly governs the spread of the disease over time, and then continue with its spatial resolution. All simulations were implemented and performed in *Octave* 5.2.0 using packages *optim* 1.6.0, *statistics* 1.4.1, *io* 2.4.13, *parallel* 3.1.3, and *splines* 1.3.3.

### Basic SIQRD model

In contrast to many existing studies that use a standard SEIR(D) model including a latency period between being infected and becoming infectious [18, 22], we focus on the difference between asymptomatic or mildly symptomatic, unknown cases that account for the major share of new infections [25, 26], and people with noticeable symptoms. We integrate this knowledge and use an SIQRD model [12, 16] that specifically distinguishes between the infectious group I, representing a measure for the estimated total number of infections, and a group Q representing known, and therefore quarantined, infections, who do not infect others anymore [14]. In our case, the transition rate *α* from I to Q describes how long it takes for an infected person to be tested/detected and put in quarantine. The remaining three groups are considered as usual, where S represents the initial state of being susceptible, R represents truly recovered, and D dead. Some fraction of I can directly recover at rate *γ*_1_ without ever being tested, overall representing the hidden infections that are never detected, while the remainder transitions to Q at rate *α*. The rate *γ*_2_ describes the rate to recover from a tested infection, while *δ* represents the rate to die from a confirmed infection (see schematic in Fig. 1). Overall, we obtain the set of equations

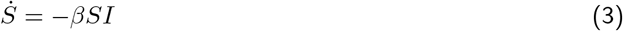

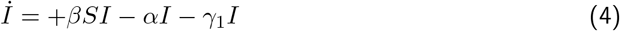

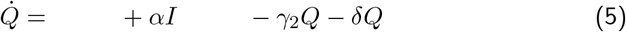

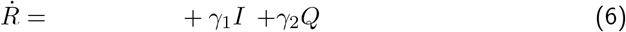

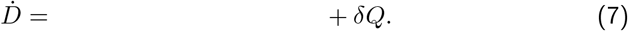

Since we are neglecting disease unrelated births and deaths, the total number of people *N* is constant, in Germany *N* ≈ 8*e*7, such that in normalized terms *S* + *I* + *Q* + *R* + *D* = 1, and therefore 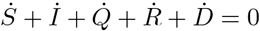.

For easier interpretation of the parameters, we introduce the dark figure *ω* = 1 + *γ*_1_*/α* and the true mortality *µ* = *δ/*(*γ*_2_ + *d*) *α/*(*α* + *γ*_1_) as given in the Results Section. Thus, we can replace the previous parameters *α* and *d* by

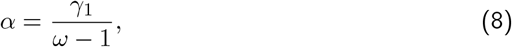

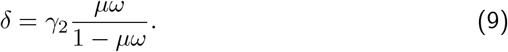

If we consider the stationary point when the pandemic has passed, we can directly relate the number of confirmed or tested infections with the estimated overall number of infections By 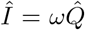, while *µ* represents the fraction of all infected people that died. The true mortality *µ* is also often referred to as infection fatality rate (IFR). Further, the mortality can also be represented by the ratio of cumulative total infections over deaths,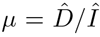. Since 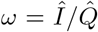, we identify the stationary relationship

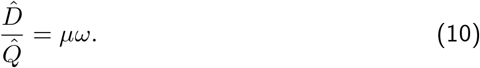

In other words, the measurable ratio *D/Q*, usually referred to as the case fatality rate (CFR), will eventually approach the number *µω*, demonstrating the inherent dependence of the two parameters. During the course of the pandemic, however, CFR will not be constant [29].

### Modeling political measures and contact restrictions

We model the effect of contact restriction and other political measures through a time-dependent infection rate *β* = *β*(*t*) by introducing three reduction factors *β*_red,*i*_ via step function 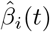, such that the effective contact rate yields

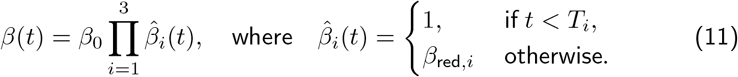

The three reduction factors *β*_red,*i*_ represent the major restrictions of 1) cancelling large events, 2) school closings, and 3) contact restrictions. We assume that each of them are constant over all of Germany, but consider their state-dependent starting date *T*_*i*_ following the respective political timeline. From preliminary investigations we observed that current data do not allow to robustly distinguish three reduction factors, manifested by parameter dependencies between *β*_red,*i*_ in our optimization scheme. Thus, we set *β*_red,2_ = 1 and used only *β*_red,3_ as a jointly fitted value to avoid over-fitting the data.

### Reproduction number

The model allows for a straight-forward estimate on the initial and effective reproduction number *R*_0_ and *R*_eff_, respectively, which are well-known in public and the general media as the number of infections originating from one infected person. Since this number is represented by the ratio between the influx and outflux of the hidden infectious group I in our model, we obtain the time-dependent upper-bound expression

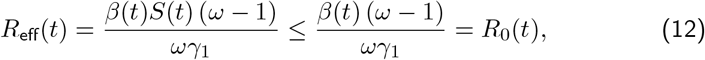

where, in relative numbers, *S* = 1 during early stages of the pandemic. Note that model parameters such as *α* and *ω* may also vary over time, and a continuous or even randomized representation of the evolution of *R*_0_(*t*) [28] may potentially better explain the imperfect data. For better readability, we drop the time-dependence of *β* and *R*_0_ in the following. As can be inferred from the expression, the uncertainty associated with the dark figure and the infection rates translate somewhat into the reproduction number, making reliable bounds on *R*_0_ difficult to obtain.

### Spatially resolved SIQRD model

In order to study the spatial dynamics of the spreading disease, we consider a network model on the resolution level of individual counties that allows for cross-county infections. Data analysis and preliminary simulations had shown that we require a federal-state-dependent dark figure *ω*_*i*_ and infection rate *β*_*i*_, *i* = 1, …, 16. On a discrete county level, for an overall number of *n*_*c*_ counties, the set of reparameterized coupled SIQRD network di_erential equations for each county k = 1; : : : ; n_c_ yield

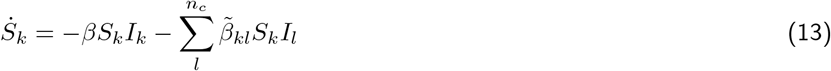

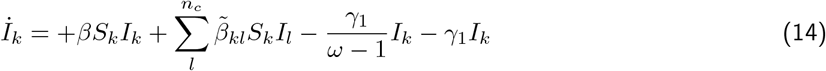

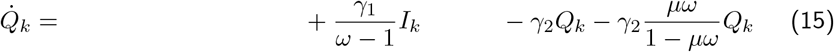

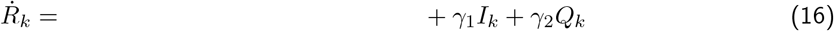

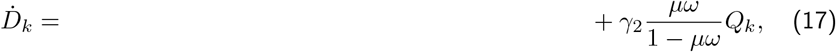

with the cross county infections

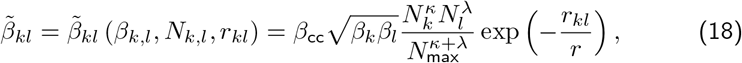

where *β*_cc,*i*_, *i* = 1, …, 16 are the state-dependent cross-county infection weights, *N*_*k*_ is the number of inhabitants in county *k, N*_*max*_ = 3*e*6 corresponds to the number of inhabitants in Germany’s largest city Berlin, and *r*_*kl*_ is the distance between counties *k* and *l*. Note that *β, ω* and *β*_cc_ are state-dependent, but not county-dependent, while all other parameters are identical for all of Germany.

The exponential cross-county infection term is adopted from the Global Epidemic And Mobility (GLEAM) model, where the expression represents commuter flows between communities *k, l*. It can be tuned by three parameters *κ, λ*, and *r* that were fit to large amounts of commuting data to globally emulate their patterns, as described in [24] (suppl. table S2). We reuse their expression in a simplified fashion and argue that cross-county infections are proportional to the commuting flow, up to the size of the infectious group at the distant county *I*_*l*_, local susceptibles *S*_*k*_ and *β*_cc,*i*_, *i* = 1, …, 16.

### Initial conditions and parameter fitting

We use data from the Robert-Koch Institute (RKI) that is available for each county in Germany over time [7]. Since RKI infection data has limited information content, we had to fix several parameters from other data describing the course of infection. Following the works of An der Heiden [9], we set the mortality to *µ* = 0.006. As described by various other works [9, 10, 37], the time to recover from a confirmed infection varies between 18 and 25 days, while milder, often undetected infections last for about 5 to 10 days. In agreement, we chose *γ*_2_ = 0.04 and *γ*_1_ = 0.067, assuming that about 50% of cases are asymptomatic [38] and undetected over a time-span of 7.5 days. Following the assumed mortality and an average time-to-death for a confirmed infection of 15 days [9], state-wise dark figures can be directly read-out from the RKI reported death toll [7] on April 25, the last day of our fit. We decided against fitting the D group over time, due to the disparate mortality across the age structure of infected people [29] which is not very well represented in SIR-type rate-based models. The age-dependent mortality was clearly visible in Germany, especially during the early stages when younger people were over-proportionally affected, with correspondingly low death rates.

For our parameter optimization, we solve the nonlinear set of ordinary differential equations (ODEs) from the start date onward in time using an ODE45 integration scheme with variable time-stepping and evaluate the daily new and cumulative infection numbers via spline interpolation.

To approach our spatially resolved county-model, we followed a cascade optimization strategy. Using state-wise identified dark figures *ω*_*i*_ and constant *µ, γ*_1_, and *γ*_2_, we first used a 16-node network model connecting each federal state to obtain a Germany-wide average *β* and reduction factors *β*_red,1_ and *β*_red,3_ by fitting the cumulative data for Germany. We then considered state-wise data to fit *β*_*i*_, *i* = 1, …, 16, while keeping *β*_cc_ = 1. As initial values, we set the number of confirmed infections on our start date March 2 as the size of *Q*_0_. To obtain an appropriate size of *I*_0_, we estimate the change rate of Q on our start date via an exponential function and then exploit 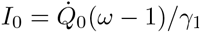.

We fit the cumulative number of confirmed infections from the RKI for the time period from March 2 until April 25 with the cumulative entries in our Q group, normalized by the maximum number of RKI infections. This is the time period during which the various shutdown measures were in place without any noticeable relaxation. On top of that, we include the change-rate of infections on our last day April 25 into the residual vector. Note that cumulative infections at time *T* reported by the RKI correspond to the integrated influx 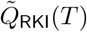 into the Q group of our SIQRD model, such that the fitted expression is obtained via

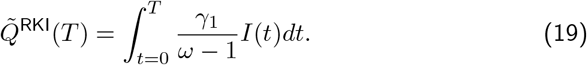

Finally, we increased the resolution to full county level, amounting to a network of 401 nodes. We used a gradient-descent algorithm to iteratively fit state-wise cross-county weights *β*_cc,*i*_, *i* = 1, …, 16 to re-balance the changed influence of the larger network, while keeping the previously determined state-wise *β*_*i*_ fixed.

The high-resolution network model brings with it the challenge for spatially consistent initial conditions. Thus, we selected the distribution of initial infections according to the RKI database on March 16, scaling down the overall number of infections to the number reported on our starting date, March 2. The ratio between Q and I was computed as before.

For the Heinsberg/Ischgl simulations, we started out with an I group of 30 times the number of inhabitants in Ischgl, amounting to *I*_0_ = 1617 30 = 48510, to represent the major tourist flow through the town and returnees from other ski resorts in Austria and Italy. In Heinsberg, we set 10% of the population in the I group, i.e., *I*_0_ = 4195, corresponding to about 65% of the population found infected in [30]. In Ischgl, we further chose *β*_cc_ equal to the highest found one in a German state (Brandenburg) to initiate the spreading, and chose Bavaria’s *β* value due to its spatial proximity.

The entire fitting procedure, except for obtaining the final cross-county weights *β*_cc,*i*_, was done using a particle swarm optimization (PSO) scheme. PSO is a meta-heuristic inspired by the behavior of natural animal swarms. It uniformly initializes a swarm of particles in a multidimensional search space, such that the objective function is evaluated at the current position of each particle. Particles communicate their best position amongst each other. Thereby, individual particle direction and speed are updated depending on their own and the overall best position in search space found up to this point. This way, the swarm broadly covers the bounded search space [39] and likely converges to a global optimum, while exploring many local minima along the way [40]. The scheme balances broad coverage with fast convergence and provides valuable information on explored samples.

### Statistical analysis

To validate the model, we evaluated the temporal and spatial correlation between model predictions and RKI data by computing the Pearson correlation coefficient *r*_*P*_, the coefficient of determination 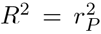 and the corresponding *p*-value to assess statistical significance via the function [*r*_*P*_, *p*] = *corrcoeff* (…) in *Octave* 5.2.0.

## Data Availability

Data on the spread of COVID-19 in Germany is publicly available from the respective resources as cited in the paper. We curated county-wise infection data over time from the RKI dashboard [7]. Data on performed tests is publicly available from the Antibiotika Resistenz Surveillance (ARS) instute of the RKI [31]. Simulation scripts are available from the corresponding author at reasonable request.
7. Robert Koch Institute. COVID-19-Dashboard, last accessed May 28, 2020.
31. Robert Koch Institute. Laborbasierte Surveillance von SARS-CoV-2 - weekly report, last accessed June 4, 2020.

https://experience.arcgis.com/experience/478220a4c454480e823b17327b2bf1d4/page/page_1/

https://ars.rki.de/Content/COVID19/Main.aspx

## Acknowledgments

We thank Dr. Lukas Pflug and the entire COVID-19 modeling group at FAU for valuable discussions and feedback on this work.

## Author contributions

PS, AK, CB, DB, and SB designed the study and developed the model. AK, CB, DB, and DL implemented the code and generated the results. MK implemented the PSO. SN acquired the data. DB performed analytical analyses. AK, CB, DL, and SB designed the figures. DB and SB wrote the original draft and supervised the model implementation. All authors have discussed the results, and reviewed and approved the final manuscript.

## Data availability

Data on the spread of COVID-19 in Germany is publicly available from the respective resources as cited in the paper. We curated county-wise infection data over time from the RKI dashboard [7]. Data on performed tests is publicly available from the Antibiotika Resistenz Surveillance (ARS) instute of the RKI [31]. Simulation scripts are available from the corresponding author at reasonable request.

## Supplementary Information

### Supplementary tables

**Table S1:**
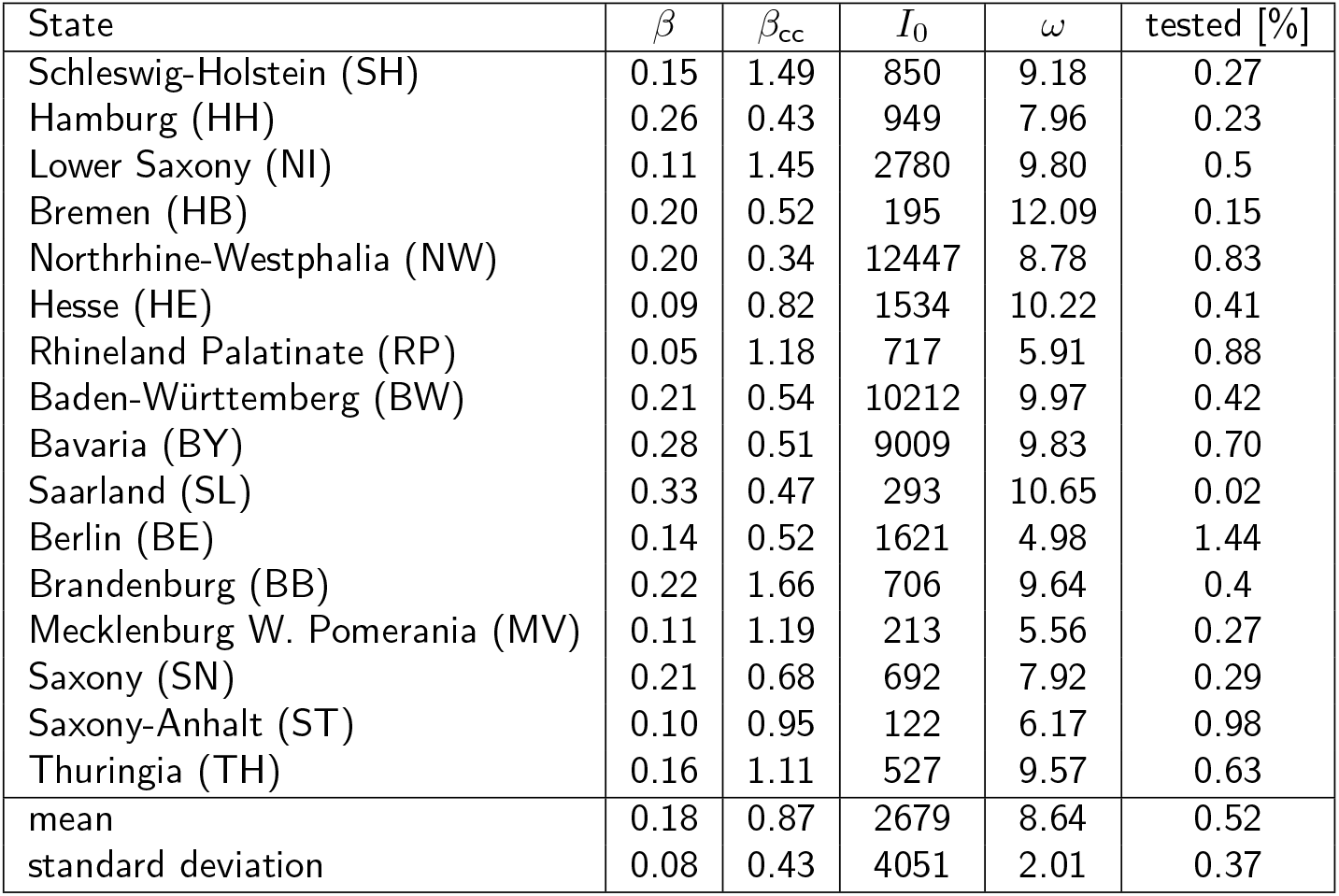
Optimized state-wise parameters for the spatially resolved SIQRD model, and percentage of population tested by April 24 [31].

| State | $\beta$ | $\beta_{cc}$ | $I_0$ | $\omega$ | tested [%] |
| --- | --- | --- | --- | --- | --- |
| Schleswig-Holstein (SH) | 0.15 | 1.49 | 850 | 9.18 | 0.27 |
| Hamburg (HH) | 0.26 | 0.43 | 949 | 7.96 | 0.23 |
| Lower Saxony (NI) | 0.11 | 1.45 | 2780 | 9.80 | 0.5 |
| Bremen (HB) | 0.20 | 0.52 | 195 | 12.09 | 0.15 |
| Northrhine-Westphalia (NW) | 0.20 | 0.34 | 12447 | 8.78 | 0.83 |
| Hesse (HE) | 0.09 | 0.82 | 1534 | 10.22 | 0.41 |
| Rhineland Palatinate (RP) | 0.05 | 1.18 | 717 | 5.91 | 0.88 |
| Baden-Württemberg (BW) | 0.21 | 0.54 | 10212 | 9.97 | 0.42 |
| Bavaria (BY) | 0.28 | 0.51 | 9009 | 9.83 | 0.70 |
| Saarland (SL) | 0.33 | 0.47 | 293 | 10.65 | 0.02 |
| Berlin (BE) | 0.14 | 0.52 | 1621 | 4.98 | 1.44 |
| Brandenburg (BB) | 0.22 | 1.66 | 706 | 9.64 | 0.4 |
| Mecklenburg W. Pomerania (MV) | 0.11 | 1.19 | 213 | 5.56 | 0.27 |
| Saxony (SN) | 0.21 | 0.68 | 692 | 7.92 | 0.29 |
| Saxony-Anhalt (ST) | 0.10 | 0.95 | 122 | 6.17 | 0.98 |
| Thuringia (TH) | 0.16 | 1.11 | 527 | 9.57 | 0.63 |
| mean | 0.18 | 0.87 | 2679 | 8.64 | 0.52 |
| standard deviation | 0.08 | 0.43 | 4051 | 2.01 | 0.37 |

**Table S2:**
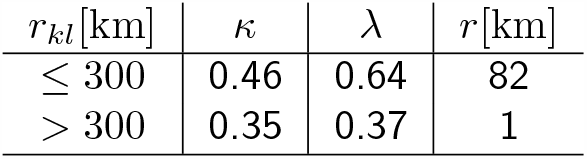
Parameters for the network mobility model as derived in [24].

### Supplementary movies

Movie S1. Spatio-temporal prediction of COVID-19 outbreak dynamics in Germany at county level from March 2 until June 2.

